# Impact of COVID-19 pandemic on severity of illness and resources required during intensive care in the greater New York City area

**DOI:** 10.1101/2020.04.08.20058180

**Authors:** Omar Badawi, Xinggang Liu, Iris Berman, Pamela J Amelung, Martin E. Doerfler, Saurabh Chandra

## Abstract

**Objective:** Describe the changes in patient population, bed occupancy, severity of illness and ventilator requirements across a large health system in the greater New York City area during the pandemic response in comparison with the 2019 baseline.

**Design:** Observational, descriptive study of ICUs monitored by a tele-ICU system across Northwell Health. Inclusion criteria: All patients admitted to Northwell Health tele-ICUs during 2019 and between March 23, 2020 and April 6, 2020.

**Exposure:** A data extract was developed to collect data every hour for each ICU bed in the Northwell tele-critical care program as a quality reporting initiative to understand ICU capacity and resource utilization. A similar extract was developed for each hour of 2019.

**Main Outcomes and Measures:** Average of any given hour during the pre-COVID-19 and pandemic periods for the following metrics: proportion of beds occupied, proportion of ventilated patients, severity of illness (measured by the ICU Discharge Readiness Score (DRS)), and length of stay (LOS).

**Results:** Hourly analysis of data from 186 ICU beds from 14 ICUs and 9 hospitals were included, representing 10,714 patients in 2019 and 465 patients between March 23 and April 6, 2020. Average hourly occupancy increased from 64% to 78%, while the proportion of patients invasively ventilated increased from 33.9% to 84.2%. Median DRS (severity of illness score) increased from 1.08 (IQR: 0.24-6.98) to 39.38 (IQR: 12.00-71.28). Proportion of patients with Hispanic ethnicity doubled (7.8% to 16.6%; p<0.01) and proportion of female patients decreased from 46.3% to 32.9% (p<0.01).

**Conclusions and Relevance:** In addition to the expected increase in ICU occupancy and ventilator requirements, this large group of ICUs in midst of the COVID-19 epidemic are faced with managing a cohort of ICU patients with a dramatically higher severity of illness than their typical census.

## Introduction

Since its emergence in December 2019 the novel coronavirus SARS-CoV-2 has become a major public threat worldwide. Immediate access to reliable up-to-date scientific information is critically important and can potentially impact outcomes for both individuals and countries alike. Experts debate the impact this pandemic will have on medical resources, such as ventilators and ICU beds, but there is little shared objective data across hospitals and primarily only anecdotal evidence exists to understand the stress our critical care system is currently under^1,2^.

Tele-critical care networks provide a unique ability to understand current data across large networks of hospitals. The Philips eICU program aggregates data from the community of eICUs for routine benchmarking, data analytics and collaborative research^3,4^. This system provides unique access to granular, clinical data in real time, including ventilator use and dynamic severity of illness scoring^5-7^.

The goal of this brief report is to describe observed changes to critical care in the face of the COVID-19 pandemic at a large network of 9 hospitals in the greater New York City (NYC) area.

## Methods

Northwell Health is a large health system in the greater NYC area using a tele-ICU system to provide supplementary coverage for many of its ICUs. Data integration occurs from the electronic health record and vital sign monitoring devices via HL7 interfaces. Additionally, the ICU admission diagnosis and other care plans are captured directly in the remote monitoring software. On March 23, 2020 a routine query to extract data related to unit type, patient demographics, mechanical ventilation use, severity of illness (DRS), and outcomes was installed to obtain a de-identified snapshot of each monitored ICU bed and transferred securely to Philips Healthcare for quality improvement analytics and reporting. For comparison, a query was run on a historical cohort from the same health system across all of 2019. Due to the prior reporting process requiring patients be discharged from the hospital for five days prior to analysis, we were limited in our ability to describe data in the weeks immediately prior to installing the prospective query.

In order to only include ICUs using continuous monitoring, as opposed to consultative monitoring, the analysis was restricted to ICUs with at least five monitored beds. Due to the urgency of describing this data, we focused on describing global critical care needs, rather than those specific to COVID-19 patients given lack of precise method to identify COVID-19 patients in the existing data structure.

### Analysis

Data were separated into pre-COVID-19 (all of 2019) and pandemic periods (March 23, 2020 to April 6, 2020). Data in the weeks immediately before the implementation of prospective data extract (1/1/2020 to 3/22/2010) were discarded due to the diminished availability of data for hospitalized and recently discharged patients. DRS was validated as a severity of illness score across the entire cohort of discharged patients by calculating the area under the receiver operating characteristic (AUROC) curve for admission, mean, median and last DRS with ICU mortality.

To describe the time-varying statistics of patients in the included ICUs, hourly patient data were summarized and compared between the pre-COVID-19 and the pandemic periods: time from ICU/hospital admission to observation time, DRS, percentage of invasive and noninvasive ventilation. Daily occupancy was defined as the daily median proportion of occupied beds of each unit in the prior 24 hours.

Patient-level statistics such as age, gender and ethnicity were also compared between the pre-and post-COVID-19 periods. Admission DRS (highest DRS score within 24 hours of ICU admission) and discharge DRS were evaluated among the discharged patients. Means and standard deviations were compared using the two-sample t-test and repeated measures ANOVA when independence of samples cannot be assumed. Skewed continuous variables were compared using the Wilcoxon rank-sum test. Categorical variables were compared using the Chi-Square test.

The expected mortality for the observed severity of illness scoring is provided through a secondary analysis of a previously reported study validating hourly assessment of 561,478 patients^7^.

## Results

These data represent 186 beds from 14 ICUs in 9 hospitals. There were 10,714 patient unit stays in the pre-COVID-19 phase and 465 in the pandemic data collection period. Table 1 describes the cohort with increases in proportion of patients of Hispanic origin and male gender. Percentages of patients with a primary admission diagnosis of either viral pneumonia or pulmonary sepsis increased from 8.2% to at least 55.1%.

**Table 1.** **Patient characteristics comparing the pre-COVID-19 and pandemic periods**

|  | **Pandemic Period\*** | **Pre-COVID-19 Period** |
| --- | --- | --- |
| **All patients** |  |  |
| **N** | 465 | 10714 |
| **Age, years (mean (SD))** | 63.72 (14.34) | 66.32 (17.13) |
| **Gender (%)** |  |  |
| Female | 153 (32.9) | 4956 (46.3) |
| Male | 312 (67.1) | 5629 (52.5) |
| Other/Unknown | 0 (0.0) | 129 (1.2) |
| **Ethnicity (%)** |  |  |
| African American | 96 (20.6) | 1777 (16.6) |
| Asian | 25 (5.4) | 602 (5.6) |
| Caucasian | 167 (35.9) | 5879 (54.9) |
| Hispanic | 77 (16.6) | 835 (7.8) |
| Native American | 3 (0.6) | 34 (0.3) |
| Missing | 0 (0.0) | 138 (1.3) |
| Other/Unknown | 97 (20.9) | 1449 (13.5) |
| **ICU admission source (%)** |  |  |
| Acute Care/Floor | 94 (20.2) | 1058 (9.9) |
| Direct Admit | 7 (1.5) | 260 (2.4) |
| Emergency Department | 220 (47.3) | 5369 (50.1) |
| ICU | 24 (5.2) | 453 (4.2) |
| Missing | 14 (3.0) | 157 (1.5) |
| Observation | 0 (0.0) | 1 (0.0) |
| Operating Room | 11 (2.4) | 1314 (12.3) |
| Other | 0 (0.0) | 4 (0.0) |
| Other Hospital | 9 (1.9) | 183 (1.7) |
| PACU | 5 (1.1) | 842 (7.9) |
| Step-Down Unit (SDU) | 81 (17.4) | 1073 (10.0) |
| **ICU type (%)** |  |  |
| Cardiac ICU | 21 (4.5) | 877 (8.2) |
| CCU-CTICU | 24 (5.2) | 358 (3.3) |
| Med-Surg ICU | 240 (51.6) | 4590 (42.8) |
| MICU | 61 (13.1) | 1959 (18.3) |
| Neuro ICU | 7 (1.5) | 119 (1.1) |
| SICU | 112 (24.1) | 2811 (26.2) |
| **Admission diagnosis grouping (%)** |  |  |
| Missing | 16 (3.4) | 410 (3.8) |
| Others | 193 (41.5) | 9427 (88.0) |
| Viral pneumonia or pulmonary sepsis | 256 (55.1) | 877 (8.2) |
| **Admission diagnosis grouping (%)** |  |  |
| Missing | 16 (3.4) | 410 (3.8) |
| Others | 113 (24.3) | 8971 (83.7) |
| Severe pulmonary disease ** | 336 (72.3) | 1333 (12.4) |

| All Completed ICU Patient Stays |  |  |
| --- | --- | --- |
| N | 311 | 10567 |
| Highest DRS within first 24 hrs of ICU (median [IQR]) | 37.46 [1.50, 71.54] | 0.94 [0.20, 7.10] |
| Last DRS before ICU discharge (median [IQR]) | 17.53 [0.76, 71.16] | 0.30 [0.09, 1.24] |
| All Surviving ICU Patient Stays |  |  |
| N | 229 | 9713 |
| Highest DRS within first 24 hrs of ICU (median [IQR]) | 13.84 [0.76, 52.09] | 0.79 [0.18, 5.47] |
| Last DRS before ICU discharge (median [IQR]) | 2.76 [0.41, 31.01] | 0.26 [0.09, 0.90] |

\*represents data collection period initiated on March 23, 2020, and through April 6, 2020. P-values were not reported as $p < 0.01$ for all comparisons.
\*\* “Severe Pulmonary Disease” was defined by the following admission diagnoses: Sepsis, pulmonary; Sepsis, unknown; Pneumonia, bacterial; Sepsis, other; Respiratory - medical, other; Effusions, pleural; Pneumonia, other; Pneumonia, viral; ARDS-adult respiratory distress syndrome, non-cardiogenic pulmonary edema;

Table 2 describes the average ICU population at any given hour during the evaluation periods. The proportion of occupied beds (64% vs. 78%; p<0.01) and patients receiving invasive ventilation at any (33.9% vs. 84.2%; p<0.01) given moment increased during the pandemic period. We also observe a dramatic increase in the severity of illness in the pandemic period. Validation of DRS as a marker of severity of illness across the entire cohort is presented in Table 3. Median DRS raised from 1.08 in pre-COVID-19 group to 39.38 in the pandemic group. Historical cohorts show that patients with a median DRS of 1.08 have an average ICU mortality of approximately 3% compared with 26% for those with a median DRS of 39.38, representing a greater than 8-fold increase in mortality risk^6^. We also observe a large increase in discharge severity of illness among ICU survivors.

**Table 2.** **ICU population characteristics comparing the pre-COVID-19 and pandemic periods**

|  | **Pandemic Period\*** | **Pre-COVID-19 Period** |
| --- | --- | --- |
| **Patient hour level statistics** |  |  |
| N, patient hours | 45339 | 890969 |
| ICU LOS, hours (median [IQR]) | 91.32 [34.63, 174.63] | 56.98 [22.27, 140.68] |
| Hospital LOS, hours (median [IQR]) | 142.85 [72.41, 243.32] | 94.62 [36.97, 244.83] |
| DRS (median [IQR]) | 39.38 [12.00, 71.28] | 1.08 [0.24, 6.98] |
| Invasive Ventilation = Yes (%) | 38184 (84.2) | 302389 (33.9) |
| Noninvasive Ventilation = Yes (%) | 520 ( 1.1) | 33534 ( 3.8) |
| **Patient hour level statistics, ventilated only** |  |  |
| N, patient hours | 38184 | 302389 |
| ICULOS, hours (median [IQR]) | 101.66 [42.23, 184.72] | 99.07 [36.35, 232.35] |
| HospitalLOS, hours (median [IQR]) | 150.49 [80.40, 245.22] | 164.53 [61.65, 381.52] |
| DRSAvg (median [IQR]) | 48.72 [23.19, 76.20] | 11.63 [3.73, 28.56] |
| **ICU daily bed occupancy** |  |  |
| Proportion of occupied beds (mean (SD)) | 0.78 (0.14) | 0.64 (0.20) |

\*represents data collection period initiated on March 23, 2020, and through April 6, 2020. P-values were not reported as $p < 0.01$ for all comparisons.

**Table 3.** **Performance (AUROC) of DRS in discriminating ICU mortality among entire cohort (n=10,878)**

| **Measure** | **AUROC (95% CI)** |
| --- | --- |
| **Highest DRS within first 24 hrs of ICU** | 0.871(0.884 - 0.897) |
| **Last DRS before ICU discharge** | 0.964(0.969 - 0.973) |
| **Mean DRS within the ICU stay** | 0.917(0.953 - 0.988) |
| **Median DRS within the ICU stay** | 0.919(0.953 - 0.987) |

## Discussion

At this time in the COVID-19 pandemic, transparency and sharing of information are crucial to risk assessments and preparedness. This is the first study to describe changes in occupancy, severity of illness and ventilator use during the COVID-19 pandemic in NYC area.

These data provide an objective view into what previously has been described only anecdotally. Although our definition for bed occupancy is intended to reflect occupied beds at any given time, the relative increase over baseline may be more interpretable given the novelty of the metric. Most notably is a dramatic increase in severity of illness during the COVID-19 timeframe. Use of invasive ventilation increased during the COVID-19 pandemic time period consistent with the known clinical course of rapidly developing respiratory failure^8-10^. With an average of 84.2% of patients receiving invasive ventilation at any given moment, this reflects a major challenge in providing care.

A change in ethnic demographics, perhaps indicating greater spread of disease in Hispanic populations, was observed. A similar finding has been noted in the African American population in Michigan^11^ and could reflect differing abilities for populations to socially distance, perhaps related to socioeconomic factors.

There are several limitations worth considering. Due to expansion of available beds for treating patients during this pandemic, these data do not reflect the entire population of ICU patients, but instead reflect patients monitored using tele-critical care. It is clear ICU patients during the pandemic are dramatically sicker than those in the prior year, possibly reflecting preferential triage to monitored units. As a supplementary model of care, it’s possible the reliability of certain data elements are suboptimal, especially amid the pandemic response though we observe low rates of missing data.

These data provide insights into the forthcoming challenges for regions on track to experience the next outbreak, which will be important for critical care management and resource planning.

## Data Availability

Data is not currently available to the public.

## Acknowledgements

We would like to thank Donna Decker, Colin McKenna, Ashley Vernon, Kurt Aglidjian and Brian Gottfried for their work to rapidly develop and deploy a system for tracking these data. Conflicts of interest include: OB, XL, and PA are all employees of Philips Healthcare.

